# Reducing Racial Disparities in Hypertension Control using a Multicomponent, Equity-Centered Approach

**DOI:** 10.1101/2024.10.28.24316324

**Authors:** Susan Mwikali Kioko, Christina Council, Cecilia Tomori

## Abstract

**Introduction:** Black Americans have the highest prevalence of hypertension among all racial or ethnic groups in the United States.^1^ They are 40% more likely to have uncontrolled blood pressure^2^ and are five times more likely to die from hypertension compared to non-Hispanic whites.^3,4^ Experiences of discrimination in healthcare, clinician and institutional bias, and socioeconomic and environmental inequities driven by structural racism contribute to uncontrolled hypertension in this population.^3,5–8^ Multilevel, multicomponent interventions have effectively improved blood pressure control among Black Americans but remain inadequately implemented in the clinical setting. An integrated nursing/public health quality improvement study was designed to address this gap between evidence and integration into clinical practice.

**Methods:** Using a one group pre/posttest design, we examined the effect of an innovative, evidence-based 12-week intervention on blood pressure among Black Americans with uncontrolled hypertension aged 18 and older in the primary care setting. Intervention components included remote blood pressure monitoring, weekly phone coaching with culturally congruent care, medication intensification, and a standardized hypertension protocol.

**Results:** The average age of the participants (n=35) was 64 years, and two thirds (n=23) were female (66%). The mean difference in systolic blood pressure from pre to post intervention decreased significantly (M=23, SD=14.0), *t*(34)= –9.7, p < .001. A significant reduction in the mean difference in diastolic blood pressure from pre to post intervention was also observed (M=11, SD=11.8), *t*(34)= –5.5, p < .001. At 12 weeks, 87% of participants had achieved blood pressure control. The intervention also improved medication adherence and hypertension knowledge (p <.001).

**Conclusion:** A multicomponent, culturally congruent quality improvement intervention significantly improved blood pressure among Black Americans.

**Health Equity Implications:** Scaled up implementation of equity-centered, culturally congruent approaches are needed to reduce racial disparities in hypertension control.

## Introduction

In the United States, Black Americans have the highest prevalence of hypertension among all racial or ethnic groups.^1^ They are 40% more likely to have uncontrolled blood pressure compared to non-Hispanic whites despite comparable hypertension awareness and treatment rates.^2,9^ Black Americans develop hypertension at an earlier age, at greater severity, and with faster disease progression than their counterparts.^10^ They are five times more likely to die from hypertension than non-Hispanic whites,^3,4^ experiencing the highest hypertension-related morbidity and premature mortality among all racial/ethnic groups.^11^

Multiple factors rooted in systemic racism are responsible for the racial disparity in hypertension prevalence and control. Individual-level factors, such as treatment adherence, are informed by health beliefs, health literacy, and experiences of discrimination in health care.^7,11^ Practice and provider-level factors, including implicit bias, influence clinical decision-making and treatment intensification.^3,12,7^ Overarching factors such as structural racism shape the socioeconomic and environmental milieu that structure the risk for hypertension and poor blood pressure control, including the affordability of medication, availability and cost of healthy food, access to recreational spaces and exposure to psychosocial and behavioral risk factors that affect hypertension control.^3,5–8^

Advancing health equity is a national priority outlined in the Framework for Health Equity,^13^ and the Healthy People 2030 Framework.^14^ Equity-based recommendations include evidence-based quality improvement initiatives that address social risk factors and gaps in health outcomes;^13^targeted interventions to improve blood pressure control among racial/ethnic minorities;^4,14^ and investing in populations that experience significant disparities in health.^13^ Previous studies have successfully utilized multicomponent interventions that combine blood pressure monitoring, drug therapy, hypertension lifestyle counseling and care coordination with community health workers (CHW) and pharmacists for hypertension management among Black Americans.^18–24^ This approach is not utilized in most primary care settings where the standard of care continues to emphasize drug therapy without systems of support for lifestyle counseling and follow-up. Dominant approaches fail to provide culturally congruent care and address health-related social needs when delivering hypertension care.^18,25^

The purpose of this study was to draw on existing evidence to develop, implement and evaluate a 12-week multilevel, multicomponent evidence-based intervention on blood pressure control among hypertensive Black Americans in the primary care setting. Our aims were to evaluate the impact of the intervention on blood pressure, medication adherence and hypertension knowledge.

## Methods

### Design

A one-group pre/posttest design was used to evaluate the effectiveness of a 12-week multilevel, multicomponent intervention on blood pressure among hypertensive Black Americans in the primary care setting. Components of the intervention included daily home blood pressure monitoring (HBPM) with remote patient monitoring (RPM); 15-minute weekly phone coaching on hypertension self-management and lifestyle; 15-minute biweekly medication management based on 2-week average blood pressure value of >140/90; and a standardized hypertension protocol. Patient education was based on the American Heart Association (AHA) guidelines. Participants were given remote blood pressure monitors to use at home during the study. The primary outcome measure was blood pressure change from enrollment using an automatic blood pressure monitor.

### Setting and Sample

The project was implemented at a primary care and specialty practice in the D.C. Metro area from September 2023 through December 2023. A sample of 35 participants were recruited through provider referral and telephone outreach. Participants were eligible for the program if: 1) self-identified as African American or Black; 2) aged 18 and older; 3) diagnosed with hypertension ICD code: I10 in EHR; 4) uncontrolled hypertension as defined by BP >140/90 or >130/80 mmHg with Type 2 Diabetes; 5) English speaking; 6) active phone line; and 7) established primary care patient. Patients diagnosed with chronic kidney disease (CKD), congestive heart failure (CHF), active malignancy or pregnancy were excluded from the study. The study was approved by the Johns Hopkins School of Nursing Ethical Review Committee.

### Intervention Procedures (see Appendix A)

Pre-implementation, a multidisciplinary team developed a standardized protocol for managing elevated blood pressure when identified during an office visit (see Appendix B). The protocol reflected best practice recommendations for hypertension management. During the enrollment period, informed consent and baseline data were collected and participants were enrolled into remote blood pressure monitoring. Patients were instructed on blood pressure monitoring following standard clinical guidelines. They were instructed to measure their blood pressure at the same time every day, and to check two readings 1-2 minutes apart. Proper measurement technique and wireless transmission of blood pressure measurement was verified at the time of enrollment, with the first measurement constituting baseline systolic and diastolic blood pressure. Questionnaires were completed through self-report or a structured interview pre/post intervention and examined for completeness. Blood pressure flow charts were reviewed daily and weekly phone coaching on hypertension self-management and lifestyle, including medication adherence, low-sodium diet, physical activity, weight management, alcohol limitation, smoking cessation, stress management and communication training was completed. Blood pressure control was assessed biweekly, and treatment intensified based on 2-week average blood pressure value of >140/90. All patient encounters were recorded in the EHR and sent to the primary care provider for review.

### Measures/Instruments

Demographic data was extracted from the EHR. Pre/post intervention blood pressure was obtained from the remote blood pressure monitor log through the measurement of systolic and diastolic blood pressure using an automatic wireless blood pressure machine. Hypertension knowledge was measured using the Hypertension Knowledge Test which has a Cronbach’s α of 0.92, indicating high reliability. It is a self-reported, 20-item questionnaire with four domains: creating awareness of the disease condition, lifestyle modifications, dietary regimen, and exercise regulation. A score of 80% indicates adequate knowledge.^15^ Medication adherence was assessed through the Morisky Medication Adherence Scale (MMAS-8). The MMAS-8 has acceptable pooled estimates of reliability in meta-analysis when used in hypertension and is widely used to assess medication adherence in chronic conditions.^16^ It is a self-reported questionnaire consisting of 8 items which assess forgetfulness, symptom severity, emotional and situation aspects of medication adherence.^16^ Higher scores indicate higher medication adherence. Feasibility was assessed through adherence to daily blood pressure monitoring, weekly attendance to phone coaching sessions, program completion and provider adherence to the hypertension protocol.

### Data Analysis

Data was analyzed using the Statistical Package for the Social Sciences (SPSS) version 29.^17^ Descriptive statistics were used to analyze characteristics of the sample population, adherence to daily blood pressure measurement, phone session attendance rate, program completion and adherence to the hypertension protocol. The mean difference in pre/post intervention systolic and diastolic blood pressure, medication adherence and hypertension knowledge test scores were analyzed using a paired *t-*test.

## Results

### Demographics

All 35 participants in this study were Black American and most were female (65.7%). Their age ranged from 34-84 years with a mean age of 64 years (±13.7). Of the 35 participants enrolled, 40% (n=14) had comorbid type 2 diabetes and 85.7% (n=30) had uncontrolled hypertension as defined by BP >140/90 or >130/80 mmHg with comorbid Type 2 Diabetes. All the participants were on antihypertensive medication at enrollment. The average body mass index (BMI) in the study sample was 32.8 (± 7.1). Thirty-four participants (97.1%) were insured at the start of the study. Most participants were not working (54.3%) with 48.6% (n=17) retired and 5.7% (n=2) unemployed. Data on education level and number of years with hypertension diagnosis was incomplete in the EHR and therefore not collected (see Table 1).

**Table 1.** Baseline Characteristics of Study Participants, N=35.

| *Study Characteristics* | *n* | *%* |
| --- | --- | --- |
| Age >65 y | 20 | 57 |
| Gender |  |  |
| Women | 23 | 66 |
| Men | 12 | 34 |
| Race (Black American) | 35 | 100 |
| Blood Pressure |  |  |
| Controlled | 5 | 14 |
| Uncontrolled | 30 | 86 |
| Comorbidity |  |  |
| Diabetes mellitus | 14 | 40 |
| Obesity (BMI >30) | 20 | 57 |
| Insurance Status |  |  |
| Medicare | 20 | 57 |
| Other | 14 | 40 |
| None | 1 | 3 |
| Employment |  |  |
| Employed | 16 | 46 |
| Unemployed | 2 | 5 |
| Retired | 17 | 49 |

### Systolic and Diastolic Blood Pressure

The average systolic blood pressure pre-intervention was 144 mmHg ±11.8 and the average systolic blood pressure post-intervention was 121 mmHg ±8.7. The mean difference in systolic blood pressure from pre to post intervention decreased significantly (M=23, SD=14.0), *t*(34)= –9.7, p < .001 (see Figure 1). The average diastolic blood pressure pre-intervention was 85 mmHg ± 12.1 and the average diastolic blood pressure post-intervention was 74 mmHg ± 6.8. The mean difference in diastolic blood pressure from pre to post intervention decreased significantly (M=11, SD=11.8), *t*(34)= –5.5, p < .001 (see Figure 2). Among the participants with uncontrolled hypertension at the start of the study (n=30), 86.7% achieved blood pressure control as defined by BP <140/90 or <130/80 mmHg with comorbid Type 2 Diabetes.

**Figure 1.**
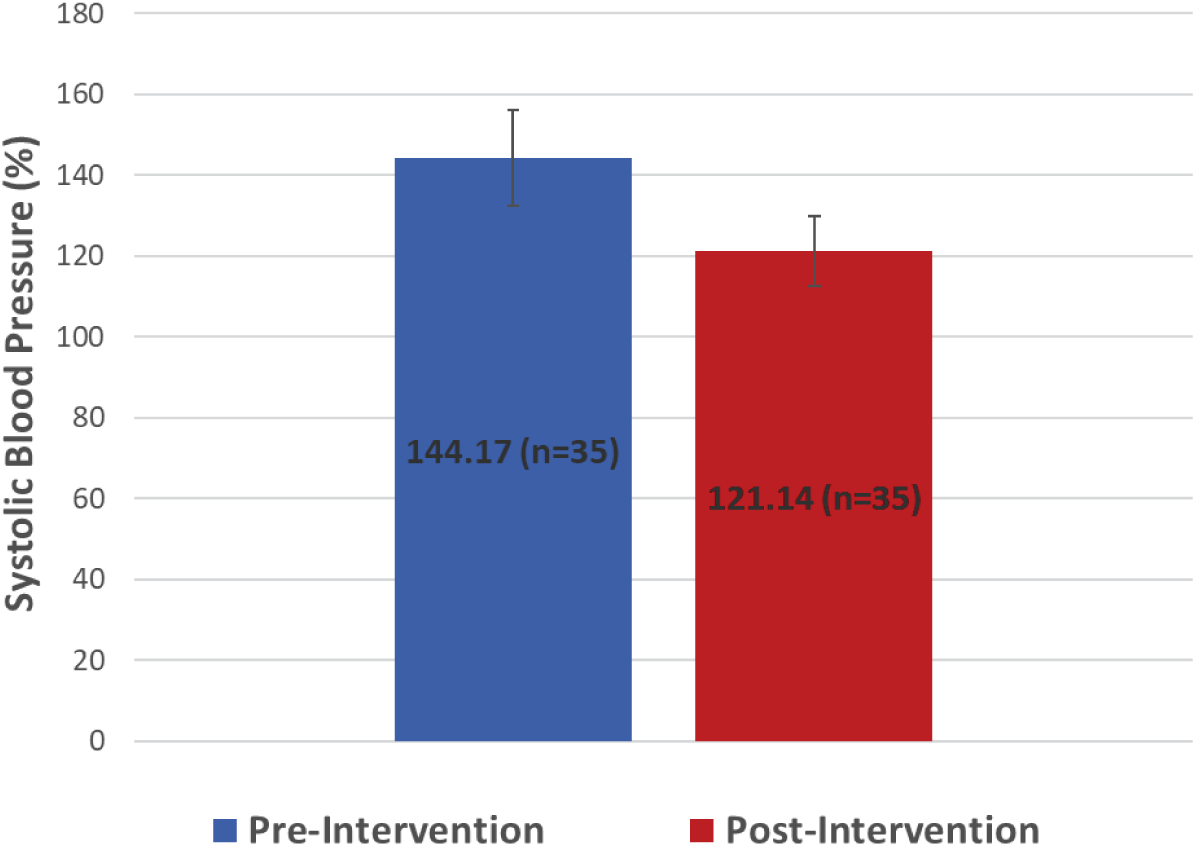
Difference in Systolic Blood Pressure Pre/postintervention, N=35. *Note*. Mean systolic blood pressure at baseline and post intervention

**Figure 2.**
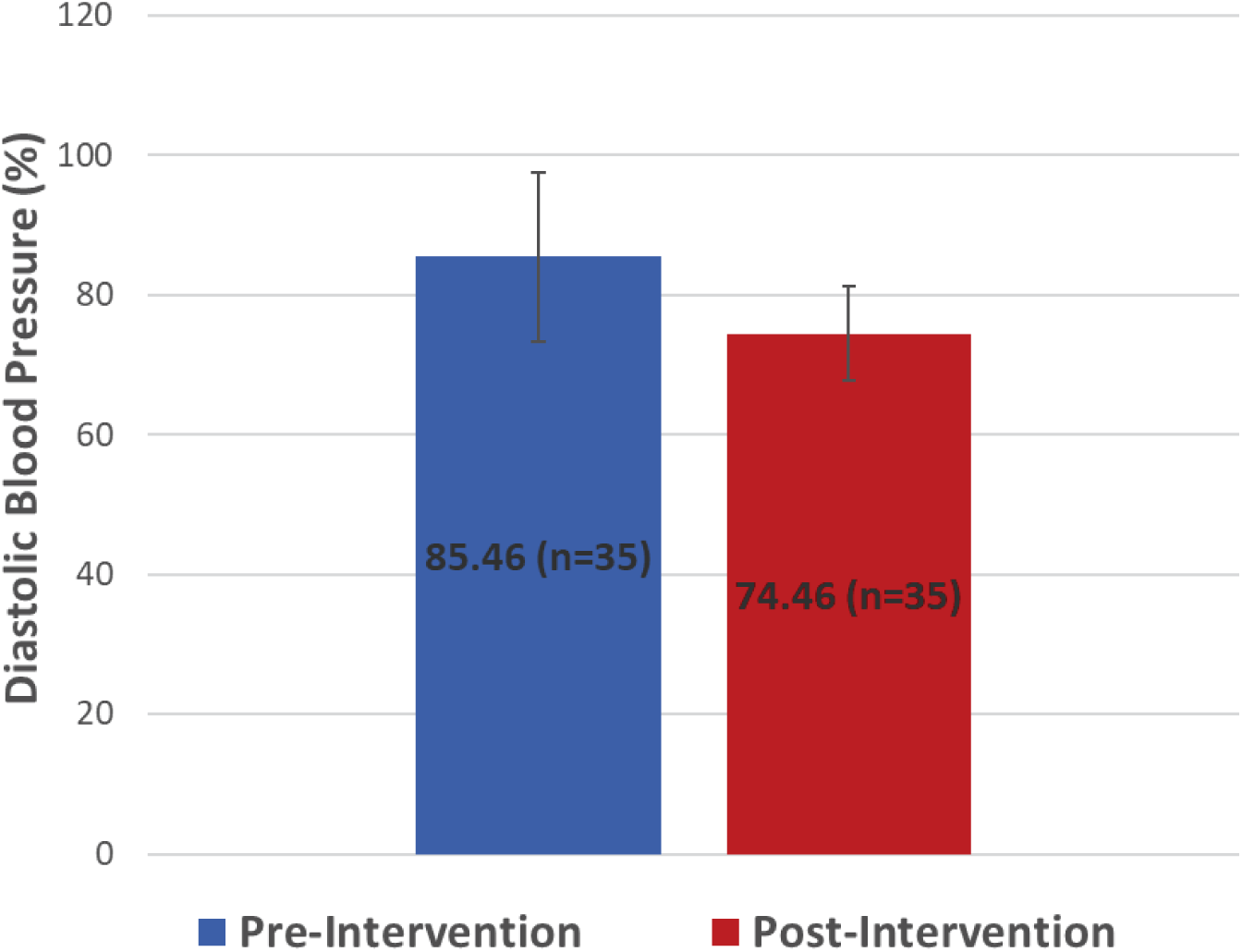
Difference in Diastolic Blood Pressure Pre/post intervention, N=35. *Note*. Mean diastolic blood pressure at baseline and post intervention

### Medication Adherence

Pre-intervention 28.6% (n=10) of participants had low medication adherence, 37.1% (n=13) had medium medication adherence and 34.3% (n=12) had high medication adherence. Post-intervention 22.9% (n=8) of participants had medium medication adherence and 77.1% (n=27) of participants had high medication adherence. No participant had low medication adherence post-intervention. The mean difference in medication adherence scores pre and post intervention improved significantly (M= 0.9, SD=1.1), *t*(34)=5.0, p < .001 (see Figure 4).

**Figure 4:**
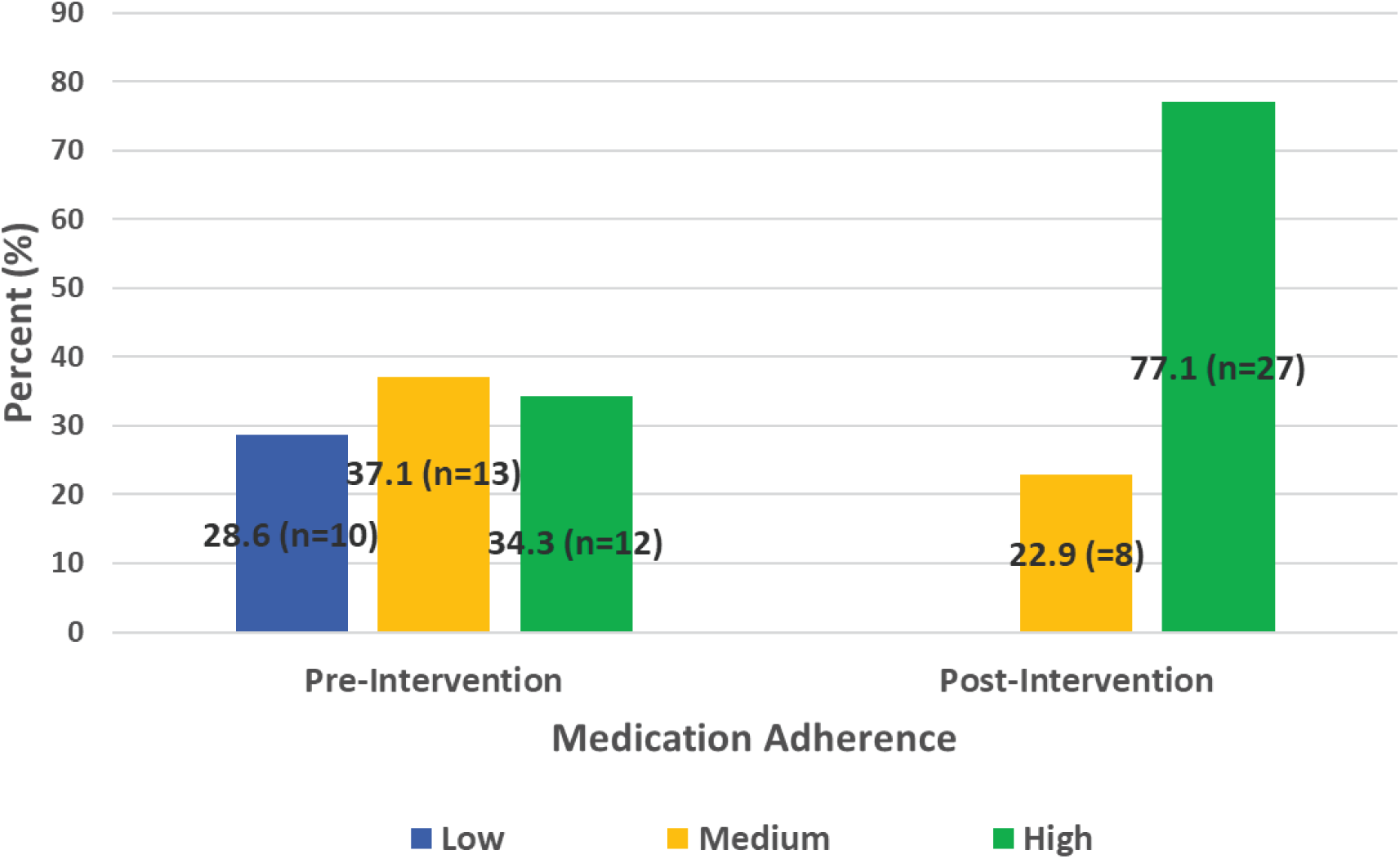
Medication Adherence Scores Pre/post Intervention, N=35

### Hypertension Knowledge

Pre-intervention, 71% (n=22) of our sample was knowledgeable as defined by a hypertension knowledge test score of 80% or greater. This improved to 97% (n=30) post-intervention. The average pre-hypertension knowledge score was 82.6% ± 9.3, and the average post-hypertension knowledge score was 94.8% ±5.6. The mean difference in hypertension knowledge test scores pre/post intervention improved significantly (M= 12.5, SD=8.2), *t*(31)=8.4, p <.001 (see Figure 5).

**Figure 5:**
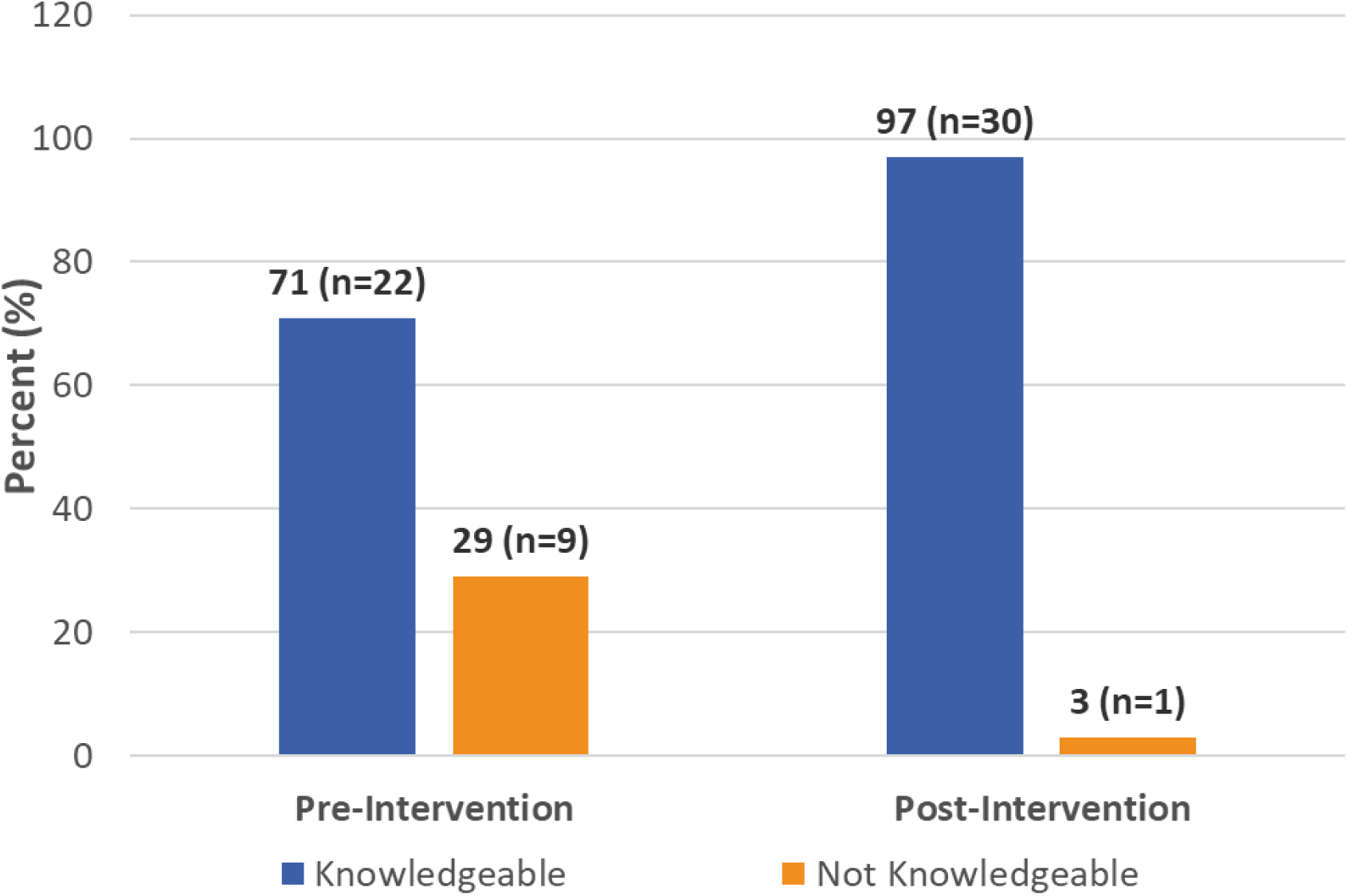
Hypertension Knowledge Test Scores Pre/postintervention, N=31

### Feasibility

All 35 participants enrolled in this hypertension disparity study completed the program. Adherence to daily blood pressure monitoring was 89.8% and the attendance rate at weekly phone coaching sessions was 94.7%. Most participants (n=31) completed post-intervention assessments except for four who did not complete the hypertension knowledge test. This resulted in 11.4% missing data. Provider adherence to the blood pressure protocol was assessed through blood pressure clinic volume, RPM enrollment, repeat blood pressure documented in the chart and the blood pressure follow-up rate. Six weeks after the hypertension practice protocol was implemented, blood pressure clinic volume was at 50% of capacity, with a consistent increase in the number of visits per week. RPM enrollment doubled and repeated blood pressures documented in the EHR increased by 9-fold. In addition, there was a 7-fold increase in the number of patients with a scheduled blood pressure follow-up. Overall adherence to the components of the blood pressure protocol remained high at 6 months except for blood pressure clinic volume; clinic visits started to decline around 3 months. Participants overwhelmingly expressed positive feedback towards the intervention throughout the study.

## Discussion

In the United States, Black Americans have the highest prevalence of hypertension,^1^ are 40% more likely to have uncontrolled blood pressure^2^ and are five times more likely to die from hypertension compared to non-Hispanic whites.^3,4^ The aim of this study was to reduce racial disparities in blood pressure control at the local level through an intervention consisting of remote blood pressure monitoring, phone coaching on hypertension lifestyle, medication intensification and a hypertension protocol.

The intervention resulted in statistically significant improvements across all outcomes, including a notable reduction of 23 mmHg in systolic blood pressure at 12 weeks. This reduction in systolic blood pressure exceeds that demonstrated in analogous studies in which systolic blood pressure reductions ranged from 9.7-21.6 mmHg over 3 to 6 months.^18–21^ Similarly, blood pressure control as defined by blood pressure <140/90 exceeded similar studies in which blood pressure control ranged from 64-81% at 4 to 6 months.^18–19,21^ Most of these studies were conducted in a community setting and with larger samples. This may have affected implementation and adherence to study protocols and ultimately study outcomes.

Significant improvements were also observed in medication adherence and hypertension knowledge. All participants were on antihypertensive medication at enrollment, however only 34% reported high medication adherence as defined by an MMAS score of 8. Self-reported medication adherence rose to 77% post intervention, with all participants reporting moderate to high medication adherence by program completion. Compared to a similar study conducted by Odemelam et al. (2020) in which phone coaching was combined with home blood pressure monitoring, our study resulted in a greater improvement in medication adherence (+43% vs +30%) despite our participants reporting less adherence to medication at baseline (66% vs 54%).^20^ This may have resulted from the combined effects and longer duration of the interventions applied in our study.

Hypertension knowledge, as defined by a hypertension knowledge test score of 80% or greater, was moderately low at baseline (71%) and increased post-intervention (97%). Compared to Odemelam et al. (2020), our intervention had a lesser effect on hypertension knowledge (+26% vs +45%) but resulted in comparable knowledge scores post-intervention (97%).^20^ This may have resulted from baseline differences in education and health literacy in our study samples, and differences in study design.

All participants completed the program. Most adhered to daily blood pressure monitoring and attended weekly phone coaching sessions. We used telehealth technology, phone calls and text messaging to avert poor attendance and completion rates noted across studies that involved on-site visits.^18, 22–24^ Provider adherence to the blood pressure protocol remained high except for blood pressure clinic follow-up. This may have resulted from increased PCP (Primary Care Physicians) follow-up and changes in clinic staffing. In addition, as RPM utilization increased, many providers were titrating medication outside of formal clinic visits, with patient communication limited to phone calls or portal messaging.

Our findings demonstrate that blood pressure, medication adherence and hypertension knowledge among Black Americans can be improved through the combination of remote blood pressure monitoring, phone coaching with culturally congruent care, medication intensification and a hypertension practice protocol. This aligns with several studies that have demonstrated the potency of a combined approach on hypertension management in this population, and offers a model for implementation in the primary care setting.^18–19,21,23,24^ Our study was not designed to assess the impact of the individual components of the intervention but suggests that weekly contact with a clinician may have engendered a sense of accountability, enhancing hypertension self-care behaviors and medication adherence. This high-touch intervention, incorporating daily reminders to check blood pressure, text messaging to reinforce treatment plans and hypertension education and weekly phone counseling, demonstrated the benefits of increased provider contact. The application of biofeedback through remote patient monitoring allowed patients to see the effect of medication and hypertension lifestyle adherence on their blood pressure in real-time, reinforcing hypertension self-care. In addition, discussing beliefs about hypertension, addressing medication aversion and side effects, and prescribing combination pills to reduce pill burden contributed to improved outcomes. Employing racial/ethnic congruence, psychoeducation and active listening during counseling fostered the development of trust, shared decision making, and self-efficacy. These adjunctive biopsychosocial interventions increased health and social support, positively impacting our outcomes.

Our intervention incorporated multiple components known to improve blood pressure control in racial/ethnic minorities, however the selected components and the application of the intervention was unique. This study embedded the intervention within a primary care practice by utilizing a nurse practitioner to extend the patient-provider relationship. Racial/ethnic and cultural congruence was employed in accord with studies demonstrating effectiveness and improved health outcomes when applied among racial/ethnic minorities.^18,25^ This helped address cultural barriers to hypertension care. Our study is one of a few to employ direct access to a health care provider through two-way text communication, to exclusively target disparities in health outcomes in the primary care setting, and to intervene at the patient, provider, and organizational level. It is also one of a few studies to examine the use of telehealth in minority populations, using technology to facilitate the implementation of a potentially complex intervention and to extend clinical care.

### Limitations

This study achieved relevant reductions in blood pressure, however the sustainability of the effects beyond the study period remains unclear. It was conducted at a single site which impacts potential scalability. Convenience sampling may have introduced sampling bias, with those who chose to participate more interested in improving their health. Our sample was older and predominately female, which may have affected study outcomes. Our small sample size limited generalizability and subgroup analyses. Although proper blood pressure measurement technique was verified at the time of enrollment, we could not assure adherence to proper technique at home which may have affected blood pressure readings. Questionnaires relied on self-report and may have been affected by language proficiency and response bias. Despite screening for eligibility through the EHR, 14.3% of participants had normal blood pressure at enrollment. During recruitment we targeted patients with blood pressures >140/90 at prior office visits, however inaccuracy of in-office readings may have led to an overestimation of blood pressure. Nevertheless, most of our sample had uncontrolled hypertension at enrollment, enabling us to demonstrate the efficacy of the intervention on blood pressure control.

## Conclusion

This study demonstrates that an intervention aimed at closing gaps in health equity is feasible and cost-effective in the primary care setting. Targeting multilevel determinants affecting blood pressure control among Black Americans improves blood pressure, medication adherence and hypertension knowledge. Future studies should explore the effect of the intervention at scale, among other racial/ethnic minority groups that experience similar disparities in blood pressure control, and on other disease conditions that commonly accompany hypertension like diabetes and obesity. Future research is also needed on the sustainability of our findings.

## Data Availability

All data referred to in the manuscript is available

## Acknowledgements

This effort would not have been possible without the support and guidance of Dr. Cecilia Tomori and Dr. Christina Council. Our deepest gratitude goes to the 35 participants who entrusted us with their care. Thank you to the providers, staff and organizational leadership who made this effort possible. Thank you to the equity researchers who paved the way for this work.

---

**Appendix A:**
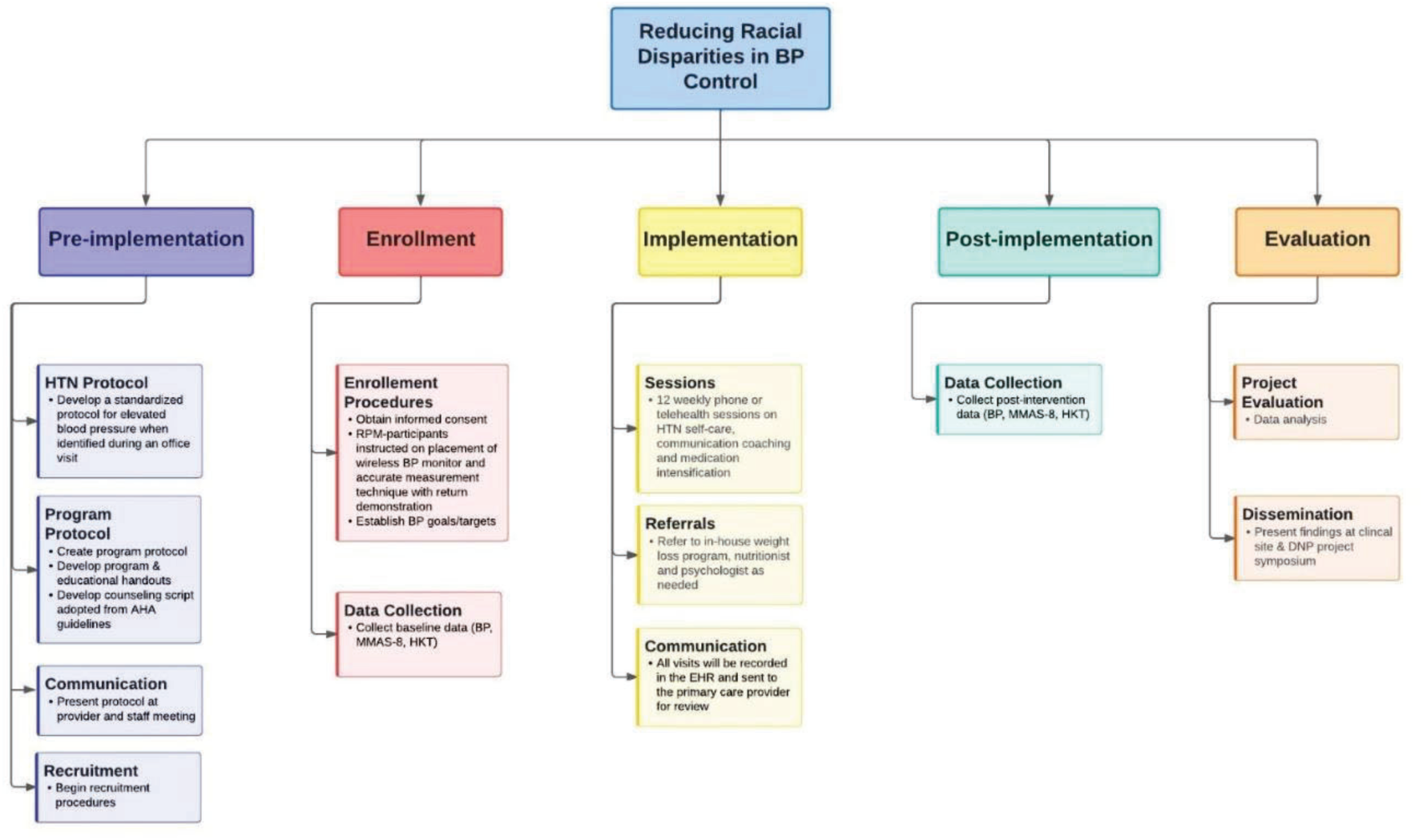
Work Breakdown Structure

**Appendix B:**
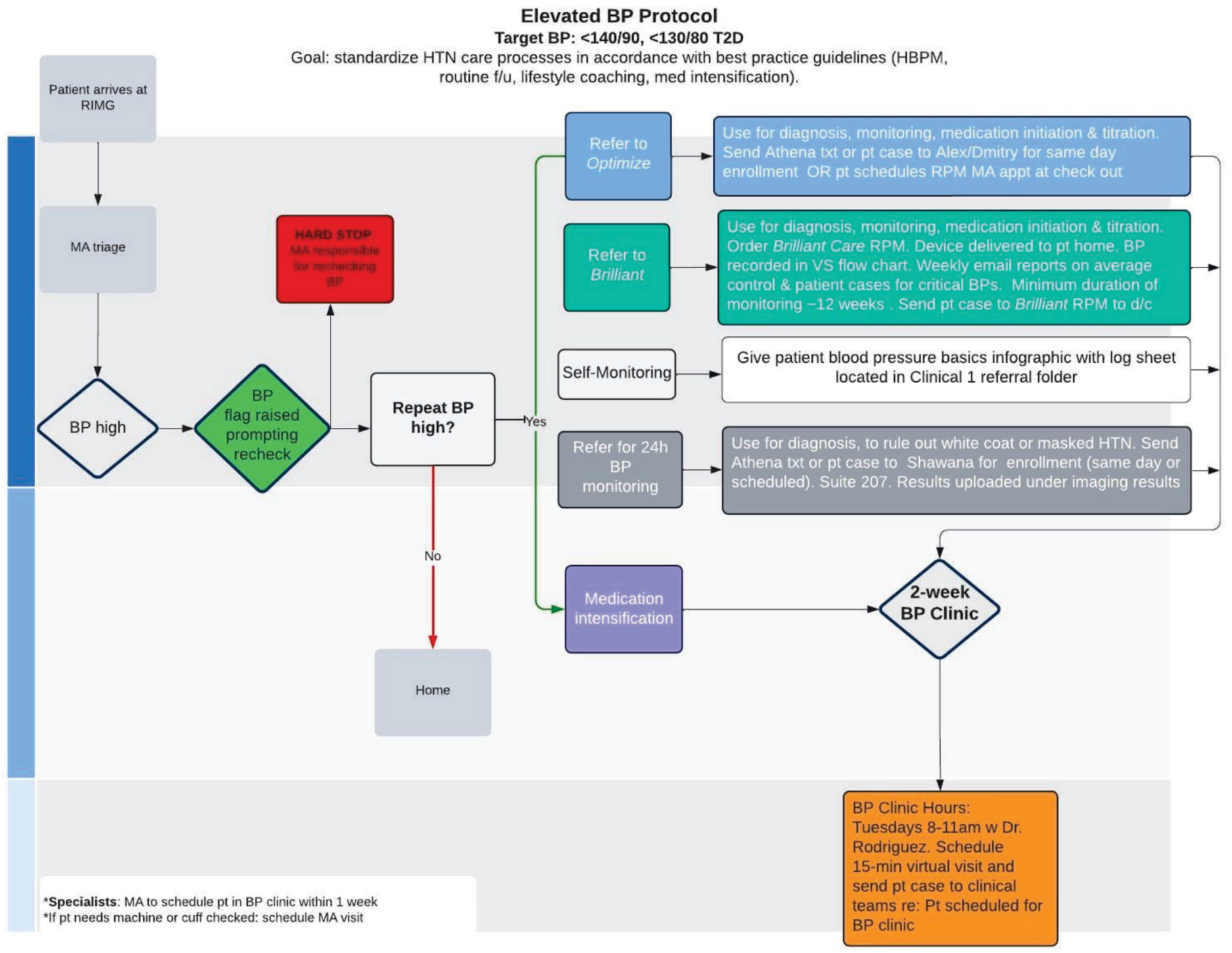
Hypertension Protocol

